# A pilot study of 0.4% povidone-iodine nasal spray to eradicate SARS-CoV-2 in the nasopharynx

**DOI:** 10.1101/2022.08.18.22278340

**Authors:** Rujipas Sirijatuphat, Amorn Leelarasamee, Thanapat Puangpet, Arunee Thitithanyanont

## Abstract

We studied the virucidal efficacy of 0.4% povidone-iodine (PVP-I) nasal spray against SARS-CoV-2 in the patients’ nasopharynx at 3 minutes and 4 hours after PVP-I exposure. We used an open-label, before and after design, single arm pilot study of adult patients with RT-PCR-confirmed COVID-19 within 24 hours. All patients received three puffs of 0.4% PVP-I nasal spray in each nostril. Nasopharyngeal (NP) swabs were collected before the PVP-I spray (baseline, left NP samples), and at 3 minutes (left and right NP samples) and 4 hours post-PVP-I spray (right NP samples). All swabs were coded to blind assessors and transported to diagnostic laboratory and tested by RT-PCR and cultured to measure the viable SARS-CoV-2 within 24 hours after collection. Fourteen patients were enrolled but viable SARS-CoV-2 was cultured from 12 patients (85.7%). The median viral titer at baseline was 3.5 log TCID_50_/mL (IQR 2.8-4.0 log TCID_50_/mL). At 3 minutes post-PVP-I spray via the left nostril, viral titers were reduced in 8 patients (66.7%). At 3 minutes post-PVP-I, the median viral titer was 3.4 log TCID_50_/mL (IQR 1.8-4.4 log TCID_50_/mL) (*P*=0.162). At 4 hours post-PVP-I spray via the right nostril, 6 of 11 patients (54.5%) had either the same or minimal change in viral titers. The median viral titer 3 minutes post-PVP-I spray was 2.7 log TCID_50_/mL (IQR 2.0-3.9 log TCID_50_/mL). Four hours post-PVP-I spray the median titer was 2.8 log TCID_50_/mL (IQR 2.2-3.9 log TCID_50_/mL) (*P*=0.704). No adverse effects of 0.4% PVP-I nasal spray were detected. We concluded that 0.4% PVP-I nasal spray demonstrated minimal virucidal efficacy at 3 minutes post-exposure. At 4 hours post-exposure, the viral titer was considerably unchanged from baseline in 10 cases. The 0.4% PVP-I nasal spray showed poor virucidal activity and is unlikely to reduce transmission of SARS-CoV-2 in prophylaxis use.

## Introduction

Coronavirus disease 2019 (COVID-19) is an acute viral respiratory infection caused by the severe acute respiratory syndrome coronavirus-2 (SARS-CoV-2) that emerged in December 2019 causing a pandemic that has killed millions [1]. The SARS-CoV-2 can persist in the upper respiratory tract of the patients at least seven days and spreads mainly via droplet and airborne transmission [2]. Patients are infectious for two to three days before the onset of symptoms, and are most contagious one to two days before the patients feel sick [3].

Povidone iodine (PVP-I) possesses rapid in-vitro virucidal activity against SARS-CoV-2 [4–7]. It can reduce SARS-CoV-2 titers by approximately 100-fold within 30 seconds after PVP-I exposure [5]. It is a broad-spectrum antiseptic and used for topical application in skin solutions and throat sprays [8–10]. PVP-I has a good safety profile and is available in Thailand. We produce 0.4% PVP-I nasal spray and use to spray aerosols into nasal cavities and nasopharynx. The aerosols containing PVP-I will lay down easily on the mucosa in the turbinate and nasopharynx and exert PVP-I virucidal activity onsite. Nasal spray used by ordinary people is easy and more convenient than nasopharynx irrigation which needs to adjust head position and the patients has to hold breathing while performing irrigation to avoid suffocation.

We hypothesized that a nasal spray of PVP-I would decrease viable virus titers in the upper airway. One practice guideline advises the use of 0.4% PVP-I nasal application and 0.5% oropharyngeal application to reduce the risk of COVID-19 transmission [11]. However, evidence of in vivo efficacy against SARS-CoV-2 in humans is very limited. Two studies reported that PVP-I administration was associated with decreased SARS-CoV-2 titers in the oral cavity of COVID-19 patients [12,13], while other studies have not demonstrated a virucidal effect of PVP-I in humans [14–16].

Therefore, we aimed to measure the speed of virucidal activity at three minutes and the sustainability of virucidal action at four hours after administration of a 0.4% PVP-I nasal spray in the patients with laboratory-confirmed COVID-19 infection. If the virucidal effect of PVP-I was rapid and at least 10 fold reduction from baseline was demonstrated in the patients’ nasopharynx, it would be beneficial to public and healthcare professionals since PVP-I nasal spray would become a simple and cost-effective measure for preventing cross-contamination and community transmission of SARS-CoV-2.

## Materials and Methods

This was an open-label, before and after study design, single-arm pilot study conducted from February 15, 2021 to March 15, 2021 at Siriraj Hospital and Samut Sakhon Hospital in Thailand and the patients were followed up for 24 hours in the hospital. The study protocol was approved by the Siriraj Institutional Review Board, Mahidol University, Thailand (COA. 408/2020), and by the Ethic Committees of Samut Sakhon Hospital (SKH REC 10/2564/V.1). Written informed consent was obtained from all study participants. This study has been registered at Thai Clinical Trials Registry under registration id. TCTR20210125002. The authors confirmed that all ongoing trials for PVP-I were registered before the patient enrollment.

Eligible patients were adults aged 18-60 years with asymptomatic or mild disease due to SARS-CoV-2 with a cycle threshold (Ct) value less than 25 of either the N gene or ORF1ab by reverse transcriptase polymerase chain reaction (RT-PCR) in the 24 hours before study enrollment. Patients with an iodine allergy or thyroid disease, pregnant or lactating women, and patients who had received antiviral agents (i.e., remdesivir and favipiravir) were excluded.

The 0.4% PVP-I was made by mixing 10% PVP-I solution with normal saline in 1:30 ratio and used as nasal spray. The following intervention was performed sequentially in all patients; 1) specimen collection by nasopharyngeal (NP) swab via the left nostril (pre-exposure specimen), 2) administration of 0.4% PVP-I nasal spray via the left and right nostrils, 3) wait 3 minutes and collect NP swab specimens through the left and right nostrils, 4) wait 4 hours and collect an NP swab specimen through the right nostril, 5) all participants were hospitalized to observe symptoms and side effects for at least 24 hours after the PVP-I nasal spray. All NP specimens were assigned numbers to blind laboratory technicians and kept in viral transport media (VTM) at 4 °C and sent to the microbiology laboratory for testing by RT-PCR and viral culture within 24 hours.

We planned to demonstrate the rapidity of 0.4% PVP-I antiviral activity by comparing the numbers of viral titer in the left side of nasopharynx (pre-exposure specimen) before and at 3 minutes after PVP-I exposure. We also compared the average viral titer in the left side of nasopharynx (pre-exposure specimens) with that of the right side of nasopharynx at 3 minutes after PVP-I exposure to provide an additional information of virucidal speed assuming that the viral titers of the left and right sides of nasopharynx were equal before the PVP-I exposure. The sustainability of antiviral activity was evaluated by comparing the viral titers in the right side of nasopharynx at 3-minutes and 4-hours after PVP-I exposure.

RNA was extracted from 200 μL of viral transport media from the NP swab samples using MagDEA® Dx kit (Precision System Science, Chiba, Japan) and SARS-CoV-2 RT-PCR was performed by amplification of SARS-CoV-2 N and ORF1ab fragments using the Detection Kit for 2019 Novel Coronavirus (2019-nCoV) RNA (PCR-Fluorescence Probing) (Da An Gene Co., Ltd. of Sun Yat-sen University, Guangdong, China), with a cycle threshold ≤40 considered positive [17]. For the viral culture test, titers were determined as the 50% tissue culture infective dose (TCID_50_) of the virus. Vero E6 cells expressing the type II transmembrane serine protease (Vero-TMPRSS2) [18] were seeded into 96-well plates and incubated with a serial dilution of the nasopharyngeal specimen. Cytopathic effects were evaluated daily until a 7-day incubation was completed.

Assuming that 70% of the pairs switch from the initial values of viable SARS-CoV-2 counts in cell culture to less than 10 fold after 3 minute of PVP-I nasal exposure and 0% from the initial value to higher viable viral count after 3 minute of PVP-I nasal exposure, and after applying continuity correction, the pilot study would require a sample size of 10 pairs to achieve a power of 80% and a two sided significance of 5% for detecting a difference of −0.70 between the discordant proportions. [19]

### Statistical analysis

Demographic data were described with descriptive statistics. Quantitative data were described with mean ± standard deviation, or median and interquartile range. Qualitative data were described in frequency (percent). The median of Ct value and viral titers were compared at each time point using the Wilcoxon signed-rank test and Mann-Whitney U test. A p value of <0.05 was considered statistically significant.

## Results

Fourteen patients were enrolled and viable culture was obtained from 12 patients (85.7%). Six patients were male (50%) with a median age 34.0 years (interquartile range (IQR) 27.5-45.0 years). Nine patients had mild disease and three patients had asymptomatic infection. The median duration from symptom onset to study enrollment was 3.0 days (IQR 1.5-4 days) (Table 1). No adverse events from PVP-I administration were observed during hospitalization.

**Table 1.**
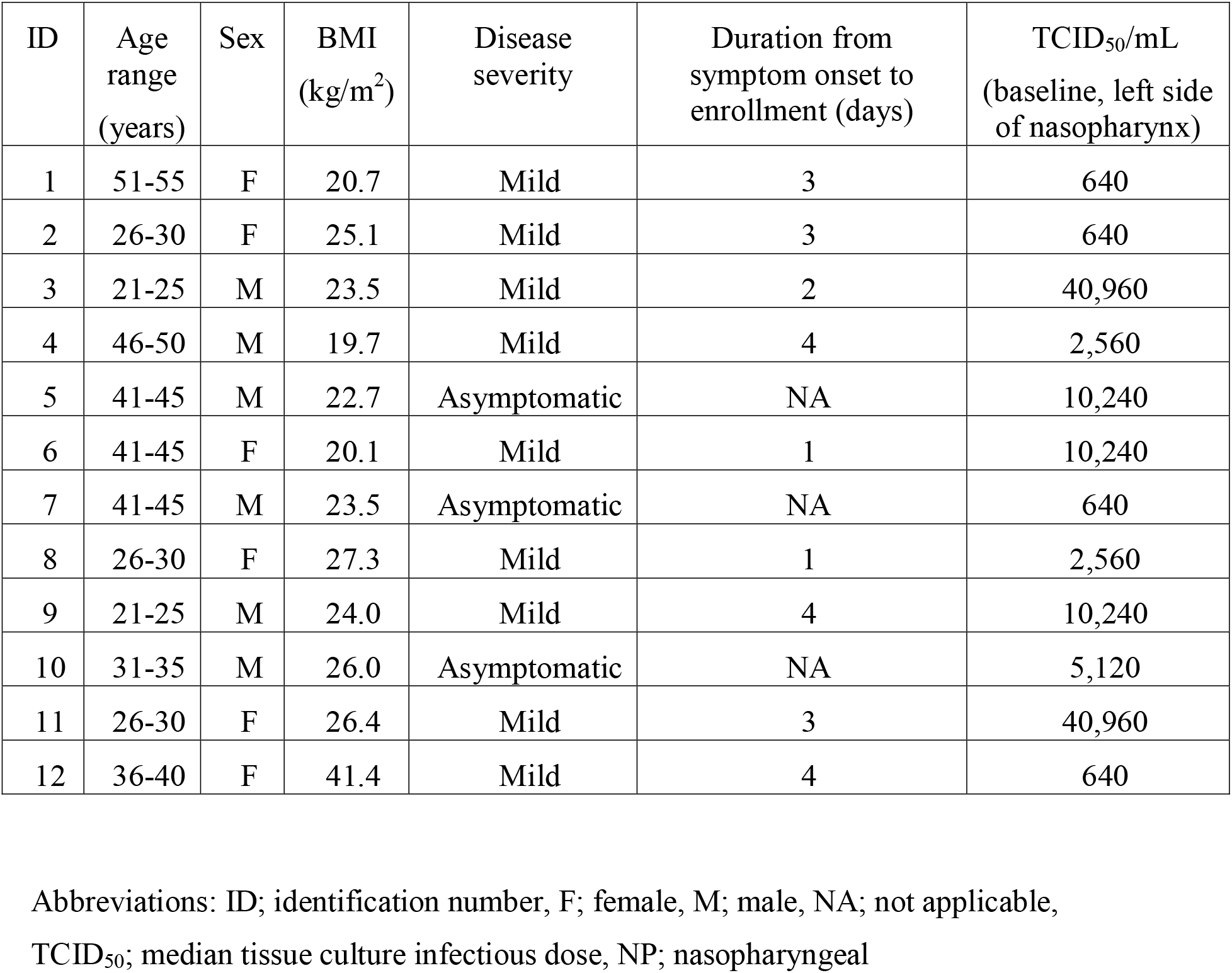
Demographic data of 12 patients with culturable NP samples

At 3 minute post-PVP-I nasal spray eight of the 12 patients (66.7%) had a reduction of viral titers. The median viral titer at baseline was 3.5 log TCID_50_/mL (IQR 2.8-4.0 log TCID_50_/mL), and the median viral titer at 3 minute post-PVP-I exposure was 3.4 log TCID_50_/mL (IQR 1.8-4.4 log TCID_50_/mL) (*P*=0.162) (Tables 2 and 3). A median value of 0.1 log reduction of viable viruses (IQR −0.05-0.23 log reduction) and 75.0% virus reduction (IQR −225.0%-93.8%) were found. The median baseline Ct value of the N gene was 19.84 (IQR 17.77-21.52), whereas the median Ct value of the N gene at 3 minute post-PVP-I was 19.49 (IQR 16.95-23.10) (*P*=0.569). The median baseline Ct value of the ORF1ab gene was 19.84 (IQR 16.82-22.22), and the median Ct value of the ORF1ab gene at 3 minute post-PVP-I was 19.17 (IQR 15.81-24.87) (*P*=0.469).

**Table 2.** Cycle threshold (Ct) values for culturable NP samples

| ID | N (baseline, Lt) | ORF1ab (baseline, Lt) | N (3 min, Lt) | ORF1ab (3 min, Lt) | N (3 min, Rt) | ORF1ab (3 min, Rt) | N (4 h, Rt) | ORF1ab (4 h, Rt) |
| --- | --- | --- | --- | --- | --- | --- | --- | --- |
| 1 | 21.96 | 23.00 | 25.42 | 26.52 | 23.44 | 24.61 | 21.93 | 22.91 |
| 2 | 22.64 | 24.18 | 23.99 | 25.45 | 20.71 | 22.34 | 24.47 | 26.12 |
| 3 | 19.71 | 21.31 | 21.54 | 23.13 | 19.32 | 20.62 | 20.41 | 21.68 |
| 4 | 20.73 | 20.14 | 20.18 | 21.47 | 19.79 | 21.16 | 19.96 | 21.29 |
| 5 | 17.73 | 19.00 | 18.54 | 19.83 | 19.74 | 21.09 | 22.23 | 23.49 |
| 6 | 18.45 | 19.53 | 19.23 | 18.08 | 18.69 | 19.87 | 18.20 | 19.32 |
| 7 | 20.40 | 22.52 | 23.62 | 26.22 | 21.49 | 23.63 | 21.27 | 23.10 |
| 8 | 19.96 | 18.46 | 19.10 | 17.72 | 17.03 | 15.72 | 18.21 | 16.91 |
| 9 | 16.19 | 15.08 | 16.42 | 15.17 | 17.36 | 16.29 | 17.94 | 16.61 |
| 10 | 17.89 | 16.27 | 16.39 | 15.07 | 15.73 | 14.50 | 16.76 | 15.39 |
| 11 | 16.30 | 15.20 | 15.40 | 14.36 | 19.41 | 18.23 | 16.91 | 15.83 |
| 12 | 21.78 | 20.38 | 19.75 | 18.51 | 24.40 | 23.30 | 24.68 | 23.47 |

Abbreviations: ID; identification number, Lt; left nasopharyngeal samples, Rt; right
nasopharyngeal samples, NP; nasopharyngeal

**Table 3.** Viral titers for culturable NP samples.

| ID | TCID<sub>50</sub>/mL<br>(baseline, Lt.) | TCID<sub>50</sub>/mL<br>(3 min, Lt.) | TCID<sub>50</sub>/mL<br>(3 min, Rt.) | TCID<sub>50</sub>/mL<br>(4h, Rt.) |
| --- | --- | --- | --- | --- |
| 1 | 640 | 10 | 80 | 160 |
| 2 | 640 | 40 | 160 | 10 |
| 3 | 40960 | 2560 | 10240 | 10240 |
| 4 | 2560 | 40960 | 320 | 640 |
| 5 | 10240 | 5120 | 320 | 160 |
| 6 | 10240 | 1280 | 10240 | 1280 |
| 7 | 640 | 160 | 40 | 320 |
| 8 | 2560 | 2560 | 640 | 640 |
| 9 | 10240 | 40960 | 640 | 5120 |
| 10 | 5120 | 40960 | 40960 | 40960 |
| 11 | 40960 | 10240 | 2560 | 20480 |
| 12 | 640 | 40 | 0 | 0 |

Abbreviations: ID; identification number, Lt; left nasopharyngeal samples, Rt; right
nasopharyngeal samples, NP; nasopharyngeal

The median viral titers of the left and right NP specimens at 3 minutes post-PVP-I exposure were 3.4 log TCID_50_/mL (IQR 1.8-4.4 log TCID_50_/mL) and 2.7 log TCID_50_/mL (IQR 2.0-3.9 log TCID_50_/mL), respectively (*P*=0.433). The median Ct value of the N gene of the left and right-side NP specimens at 3 minute post-PVP-I were 19.49 (IQR 16.95-23.10) and 19.58 (IQR 17.69-21.30), respectively (*P*=0.977). The median Ct value of the ORF1ab gene of the left and right-side NP specimens at 3 minute post-PVP-I were 19.17 (IQR 15.81-24.87) and 20.86 (IQR 16.78-23.06), respectively (*P*=0.885).

At 4 hour post-PVP-I nasal spray, six of the 11 patients (54.5%) were observed to have either the same titer or a minimal reduction of viral titers, and five patients had increased titers from the samples collected at 3 minute post-PVP-I. The median viral titer at 3 minute post-PVP-I exposure was 2.7 log TCID_50_/mL (IQR 2.0-3.9 log TCID_50_/mL), and the median viral titer at 4 hour post-PVP-I exposure was 2.8 log TCID_50_/mL (IQR 2.2-3.9 log TCID_50_/mL) (*P*=0.704). The median Ct value of the N gene at 3 minute post-PVP-I exposure was 19.58 (IQR 17.69-21.30), and the median Ct value of the N gene at 4 hour post-PVP-I exposure was 20.19 (IQR 18.01-22.16) (*P*=0.339). The median Ct value of the ORF1ab gene at 3 minute post-PVP-I exposure was 20.86 (IQR 16.78-23.06), and the median Ct value of the ORF1ab gene at 4 hour post-PVP-I was 21.49 (IQR 16.69-23.38) (*P*=0.433).

**Table 4.** Change in viral titers after 3 minute and 4 hour exposure to PVP-I as compared to the baseline value from left NP specimens.

| ID | ≈ 10 fold | Within 4 fold reduction | No change or increase |
| --- | --- | --- | --- |
|  | reduction |  |  |
| 1 | 3 min, Lt. | 3 min, Rt.; 4h, Rt. | 0 |
| 2 | 4h, Rt. | 3 min, Lt.; 3 min, Rt. | 0 |
| 3 | 0 | 3 min, Lt.; 3 min, Rt.; 4h, Rt. | 0 |
| 4 | 0 | 3 min, Rt.; 4h, Rt. | 3 min, Lt. |
| 5 | 0 | 3 min, Lt.; 3 min, Lt; 4h, Rt. | 0 |
| 6 | 0 | 3 min, Lt.; 4h, Rt. | 3 min, Lt. |
| 7 | 3 min Rt. | 3 min, Lt.;4h, Rt. |  |
| 8 | 0 | 3 min, Rt.; 4h, Rt. | 3 min, Lt. |
| 9 | 0 | 3 min, Rt.; 4h, Rt. | 3 min, Lt. |
| 10 | 0 | 0 | 3 min, Lt.; 3 min, Lt.; 4h, Rt. |
| 11 | 3 min, Rt. | 3 min, Lt; 4h, Rt. | 0 |
| 12 | 3 min, Lt.; 3 min,<br>Rt.; 4h, Rt. | 0 | 0 |

Only one case (no. 12) showed viral titers decreased approximately ten folds. The rest showed only within 1-4 fold reduction which could be due to sampling variation or weak PVP-I anti-viral activity. Five cases even had increased viral titers after PVP-I exposure. Overall, the decrease of viral titers is not rapid and not as much as shown by the in vitro test which is about 100 fold decrease 1 minute after PVP-I exposure. At 4 hour after PVP-I exposure, minimal or no change in viral titer was demonstrated. Overall, the PVP-I did not demonstrate rapid and strong virucidal activity as has been shown by the in vitro test.

## Discussion

The TCID_50_/mL of the 12 patients with culturable NP samples varied from 640 to 40,960, which was not related to either disease severity or the onset of clinical symptoms when NP swabs were taken. Because all cases recovered uneventfully, the initial values of TCID_50_/ml did not predict the treatment outcome. However, the sample size was small and we did not plan to study the relationship between the initial viral titers and disease severity or clinical outcome.

We found a median one-fold reduction of SARS-CoV-2 viral titer from nasopharyngeal swabs at 3 minute post-PVP-I exposure in the left side of nasopharynx. This was in marked contrast to in vitro studies that reported as much as a 100-fold reduction in viral titer [5]. At 4 hour post-exposure to 0.4% PVP-I nasal spray, the viral titer in the right side of nasopharynx was unchanged from baseline in 10 cases.

Although the in vitro rapid activity of PVP-I against SARS-CoV-2 has been established in several studies [5–7], evidence of in vivo activity in humans is limited. Two studies have reported that PVP-I administration into the upper aerodigestive tract of COVID-19 patients was associated with lower SARS-CoV-2 viral load and Ct values [12,13]. However, other studies have not confirmed their results [14–16]. To measure potential reductions in viral shedding, we believe that RT-PCR alone is insufficient and viral culture is needed. Using viral culture, PVP-I administration probably reduced infectious viral titers in the upper respiratory tract of COVID-19 patients in two recent studies [20,21]. Friedland P et al demonstrated that 0.5% PVP-I nasal spray was associated with a reduction of viral titers in five of the six study participants (83%) five minutes after PVP-I application [20]. Seikai T et al revealed that gargling with PVP-I was associated with significantly decreased viral titers at 60 minutes after PVP-I exposure [20].

We found that PVP-I nasal spray had poor in vivo efficacy in our study. This could be explained by several factors including insufficient concentration, improper formulation and/or amount of PVP-I, inadequate duration of exposure, inappropriate method of administration, and mucociliary clearance of PVP-I from the nasal cavity [20]. We observed that 0.4% PVP-I nasal spray did not change the Ct value of the N and ORF1ab segments and did not interfere with PCR-mediated laboratory diagnosis of COVID-19. Finally, 0.4% PVP-I administered as a nasal spray was safe and well-tolerated.

Our study had some limitations. First, this was a small, single-arm, pre- and post-exposure pilot study, therefore, the study result may differ from larger-scale studies. Second, the study participants were likely infected with the strain B.1.36.16 of SARS-CoV-2 that caused infections in Thailand during the study period [22]. Our findings may not be generalizable to other variants of SARS-CoV-2.

However, since the PVP-I has been shown to exhibit broad-spectrum, rapid virucidal activity against various respiratory viruses by the in vitro tests, the finding of our study in patients infected with SARS-CoV-2 should give warning against its use for prophylaxis in ordinary people.

### Conclusions

The 0.4% PVP-I nasal spray had minimal virucidal activity in the nasopharynx of infected patients. Hence, it is unlikely that this treatment would reduce airborne or droplet transmission of COVID-19. Other preventive measures such as vaccination, face masking, and personal protective equipment should be emphasized to limit COVID-19 transmission.

## Data Availability

All relevant data are within the manuscript and its Supporting Information files.

## Acknowledgements

The authors gratefully acknowledge Mrs. Sukanya Chanboonchuay for her assistance with project management, Dr. Paraya Assanasen (ENT physician) for making the 0.4% PVP-I nasal spray, Miss Suwimon Manopwisedjaroen, M.Sc. and Miss Chanya Srisaowakarn, M.Sc. for microbiological laboratory assistance. The authors also thank the COVID-19 patient care team of Samut Sakhon Hospital (Mr. Chaiyot Ra-ngubpai, Miss Punyanee Triamkan, Miss Laksamee Suddee, Miss Kitima Limprasert and Mrs. Walaiporn Jaiaree) for providing patient information, specimen collection, and patient observation during hospitalization.

## Declarations

### Ethical Approval

This study was approved by the Institutional Review Board of the Faculty of Medicine Siriraj Hospital, Mahidol University (COA. 408/2020), and by the ethic committees of Samut Sakhon Hospital (SKH REC 10/2564/V.1).

## References

1. COVID-19 Excess Mortality Collaborators. Estimating excess mortality due to the COVID-19 pandemic: a systematic analysis of COVID-19-related mortality, 2020-21. Lancet 2022:S0140-6736(21)02796-3.

2. Badu K, Oyebola K, Zahouli JZB, Fagbamigbe AF, de Souza DK, et al. SARS-CoV-2 Viral Shedding and Transmission Dynamics: Implications of WHO COVID-19 Discharge Guidelines. Front Med (Lausanne) 2021;8:648660.

3. Casey-Bryars M, Griffin J, McAloon C, Byrne A, Madden J, Mc Evoy D, et al. Presymptomatic transmission of SARS-CoV-2 infection: a secondary analysis using published data. BMJ Open 2021;11:e041240.

4. Pelletier JS, Miller D, Liang B, Capriotti JA. In vitro efficacy of a povidone-iodine 0.4% and dexamethasone 0.1% suspension against ocular pathogens. J Cataract Refract Surg 2011; 37: 763–766.

5. Anderson DE, Sivalingam V, Kang AEZ, Ananthanarayanan A, Arumugam H, Jenkins TM, et al. Povidone-iodine demonstrates rapid in vitro virucidal activity against SARS-CoV-2, the virus causing COVID-19 disease. Infect Dis Ther 2020; 9: 669–675.

6. Bidra AS, Pelletier JS, Westover JB, Frank S, Brown SM, Tessema B. Rapid In-Vitro Inactivation of Severe Acute Respiratory Syndrome Coronavirus 2 (SARS-CoV-2) Using Povidone-Iodine Oral Antiseptic Rinse. J Prosthodont 2020; 29: 529–533.

7. Shet M, Hong R, Igo D, Cataldo M, Bhaskar S. In vitro evaluation of the virucidal activity of different povidone-iodine formulations against murine and human coronaviruses. Infect Dis Ther 2021; 10: 2777–2790.

8. Nagatake T, Ahmed K, Oishi K. Prevention of respiratory infections by povidone-iodine gargle. Dermatology 2002; 204 Suppl 1: 32–36.

9. Ghaddara HA, Kumar JA, Cadnum JL, Ng-Wong YK, Donskey CJ. Efficacy of a povidone iodine preparation in reducing nasal methicillin-resistant Staphylococcus aureus in colonized patients. Am J Infect Control 2020; 48: 456–459.

10. Peng HM, Wang LC, Zhai JL, Weng XS, Feng B, Wang W. Effectiveness of preoperative decolonization with nasal povidone iodine in Chinese patients undergoing elective orthopedic surgery: a prospective cross-sectional study. Braz J Med Biol Res 2017; 51: e6736.

11. Krajewska Wojciechowska J, Krajewski W, Zub K, Zatoński T. Review of practical recommendations for otolaryngologists and head and neck surgeons during the COVID-19 pandemic. Auris Nasus Larynx 2020; 47: 544–558.

12. Seneviratne CJ, Balan P, Ko KKK, Udawatte NS, Lai D, Ng DHL, et al. Efficacy of commercial mouth-rinses on SARS-CoV-2 viral load in saliva: randomized control trial in Singapore. Infection 2021; 49: 305–311.

13. Elzein R, Abdel-Sater F, Fakhreddine S, Hanna PA, Feghali R, Hamad H, et al. In vivo evaluation of the virucidal efficacy of Chlorhexidine and Povidone-iodine mouthwashes against salivary SARS-CoV-2. A randomized-controlled clinical trial. J Evid Based Dent Pract 2021; 21: 101584.

14. Guenezan J, Garcia M, Strasters D, Jousselin C, Lévêque N, Frasca D, et al. Povidone iodine mouthwash, gargle, and nasal spray to reduce nasopharyngeal viral load in patients with COVID-19: A randomized clinical trial. JAMA Otolaryngol Head Neck Surg 2021; 147: 400–401.

15. Zarabanda D, Vukkadala N, Phillips KM, Qian ZJ, Mfuh KO, Hatter MJ, et al. The Effect of Povidone-Iodine Nasal Spray on Nasopharyngeal SARS-CoV-2 Viral Load: A Randomized Control Trial. Laryngoscope 2021: 10.1002/lary.29935.

16. Ferrer MD, Barrueco ÁS, Martinez-Beneyto Y, Mateos-Moreno MV, Ausina-Márquez V, García-Vázquez E, et al. Clinical evaluation of antiseptic mouth rinses to reduce salivary load of SARS-CoV-2. Sci Rep 2021; 11: 24392.

17. A R, Wang H, Wang W, Tan W. Summary of the Detection Kits for SARS-CoV-2 Approved by the National Medical Products Administration of China and Their Application for Diagnosis of COVID-19. Virol Sin 2020; 35: 699–712.

18. Sasaki M, Uemura K, Sato A, Toba S, Sanaki T, Maenaka K, et al. SARS-CoV-2 variants with mutations at the S1/S2 cleavage site are generated in vitro during propagation in TMPRSS2-deficient cells. PLoS Pathog 2021; 17: e1009233.

19. Dhand NK, Khatkar MS. (2014). Statulator: An online statistical calculator. Sample Size Calculator for Comparing Two Paired Proportions. Accessed 25 July 2022 at http://statulator.com/SampleSize/ss2PP.html

20. Friedland P, Tucker S, Goodall S, Julander J, Meddenhall M, Molloy P, et al. In vivo (human) and in vitro inactivation of SARS-CoV-2 with 0.5% povidone-iodine nasal spray. Aust J Otolaryngol 2022; 5: 2.

21. Seikai T, Takada A, Hasebe A, Kajihara M, Okuya K, Sekiguchi Yamada T, et al. Gargling with povidone iodine has a short-term inhibitory effect on SARS-CoV-2 in patients with COVID-19. J Hosp Infect 2022: S0195-6701(22)00006-8.

22. Kunno J, Supawattanabodee B, Sumanasrethakul C, Wiriyasivaj B, Kuratong S, Kaewchandee C. Comparison of Different Waves during the COVID-19 Pandemic: Retrospective Descriptive Study in Thailand. Adv Prev Med 2021; 2021: 5807056.

